# Phase Boundaries and Critical Transitions in Coupled Epidemic–Behavioral Systems

**DOI:** 10.1101/2025.10.05.25337342

**Authors:** Hsuan-Wei Lee, Vincent Cheng-Sheng Li

## Abstract

Epidemics are nonlinear adaptive processes in which pathogen spread and human behavior form a tightly coupled feedback loop. Individual decisions about protective measures create strategic interactions. These interactions can either accelerate disease spread or drive collective suppression. We introduce a theoretical lattice-based agent model that fuses SIS contagion with an evolutionary game, systematically exploring how strategy choice and infection pressure co-evolve through comprehensive parameter space analysis. Agents choose between self-isolation and normal activity based on population-wide disease prevalence and perceived costs. Agents then update strategies using a Fermi rule based on global infection prevalence and perceived costs. Infections propagate through contact-based transmission with behavior-dependent probability. We model transmission with a hierarchical probability structure where cross-infection coupling captures risk at behavioral interfaces between strategies. Comprehensive exploration of the four-dimensional parameter space reveals sharp phase transitions between cooperative and defective regimes. These transitions are governed by transmission intensity, recovery probability, risk perception, and economic pressures. A striking paradox emerges: while intense cross-infection coupling drives near-universal isolation adoption, it paradoxically sustains persistent endemic infection, demonstrating that widespread cooperation does not guarantee epidemic control. Modest changes in isolation costs or cross-infection coupling trigger complete phase inversions. This extreme sensitivity characterizes systems operating near critical points. Contact-mediated spread generates persistent spatial patterning in infection status and compartment composition. These findings establish epidemic-behavioral coupling as a fundamentally nonlinear dynamical system exhibiting critical phenomena and emergent spatial organization. Cooperation emergence does not guarantee epidemic control, revealing complex theoretical relationships between individual decision-making and collective health outcomes that require empirical validation for practical application.

## 1. Introduction

Epidemics are nonlinear systems in which a few local infections can ignite large outbreaks by exponential growth [1, 2, 3]. Minor shifts in perception or policy can suppress the same outbreaks with comparable speed [4, 5]. This extreme sensitivity exemplifies complex adaptive systems operating near critical points [6, 7]. Classical compartmental models capture parameter sensitivity yet hold human behavior fixed while pathogens spread [8, 9, 10]. Real populations adapt. People constantly weigh social freedom against infection risk. These choices create strategic interactions because individual behavior alters exposure via transmission, and changes in population prevalence feed back into strategy payoffs. The epidemic therefore becomes an evolutionary game that links biology to choice [11, 12, 13, 14]. Behavioral strategies change under selection pressures as individuals update using prevalence-dependent payoffs. These generate feedback loops that couple epidemiological progression with strategy evolution across spatial and temporal scales.

Such strategic feedbacks can reverse epidemic trajectories and reshape system behavior [15, 16, 17]. COVID-19 demonstrated how behavioral adaptations including isolation and quarantine [18, 19, 20], social distancing [21, 22], mask wearing [23, 24], and vaccination create cascades that either accelerate transmission or drive suppression [25, 26]. From an evolutionary game theory perspective, these health behaviors represent costly cooperative strategies in spatial public goods games where individual sacrifices generate collective benefits [27, 28, 29]. Vaccination confers herd immunity, mask wearing reduces transmission risk for others, and isolation eliminates infectious contacts. In our setting, the fitness of each strategy depends on population-wide infection prevalence *I*(*t*), creating frequency-dependent selection in which the attractiveness of isolation rises with prevalence. Previous work has demonstrated how evolutionary games on adaptive networks can generate complex dynamics through strategy-network coevolution [30, 31, 32], but epidemic contexts introduce additional biological constraints that fundamentally alter the evolutionary landscape [33, 34, 35, 36]. The emergence of costly cooperation depends on how contact structure shapes transmission opportunities, how easily infection crosses between behavior groups, and the strength of selection generated by infection risk. When boundaries are porous, cooperative strategies become vulnerable to exploitation by defectors who benefit from others’ protection while avoiding costs. When impermeable, cooperator clusters maintain high fitness but become evolutionarily isolated. The intermediate regime generates the richest evolutionary dynamics, where small parameter changes trigger dramatic shifts between cooperation-dominated and defection-dominated evolutionary equilibria.

Diverse approaches have advanced our understanding, including awareness-based psychology models [37] and social-network frameworks [38]. Yet many models still miss the full complexity of strategic interactions and spatial clustering. Mean-field approximations, though computationally tractable and providing important baseline insights, eliminate spatial structure that can be crucial for understanding cooperation emergence in structured populations [39, 40, 41]. Different modeling frameworks offer complementary strengths: deterministic approaches provide analytical tractability and clear mechanistic insights, while stochastic frameworks capture noise-induced transitions and variability in decision-making processes [42, 43]. Our evolutionary game-theoretic approach builds upon this rich literature by specifically focusing on spatial heterogeneity and stochastic strategy updating to understand pattern formation in epidemic-behavioral systems [44, 45]. Network-based models, while incorporating some spatial structure, typically assume fixed topologies that cannot adapt to changing behavioral dynamics [46, 47, 48]. Here we likewise use a fixed lattice but resolve spatial transmission explicitly and couple strategy payoffs to shared prevalence. A central gap is the weak treatment of cross-strategy infection, which controls whether mixed populations evolve toward cooperation or collapse into defection. Classical epidemiological models treat behavioral parameters as exogenous constants, ignoring the feedback mechanisms where epidemic state influences strategy adoption and vice versa [49, 50]. Cross-group infection sets the stability of cooperation, its invasion speed, and its resistance to shifting costs. The absence of proper cross-infection modeling is particularly problematic, as it determines whether behavioral boundaries remain permeable or become evolutionarily stable barriers that shape long-term population dynamics [51]. Without proper representation of these evolutionary game mechanisms, models cannot capture the multistability and critical phenomena characterizing real epidemic-behavioral systems, leaving policymakers with inadequate foundations for designing interventions that account for strategic behavior and evolutionary responses.

Recent work begins to address these limits using evolutionary game-theoretic models. These frameworks capture the strategic nature of health decisions during pandemics. Models incorporating voluntary isolation strategies governed by evolutionary dynamics have revealed recurrent infection waves driven by population awareness cycles, where reduced disease perception leads to cooperation breakdown and subsequent epidemic resurgence [52]. These studies demonstrate that individual risk perception creates complex feedback loops with collective epidemic outcomes, generating oscillatory dynamics where successful disease suppression paradoxically undermines the very behaviors that achieved it [53, 54]. Investigations of vaccination behavior through game-theoretic lenses have identified critical bifurcation points where small changes in perceived costs trigger dramatic shifts in population coverage, revealing the fragility of herd immunity under voluntary compliance [55, 56, 57]. Studies of mask-wearing behaviors have uncovered the role of social conformity and network topology in sustaining protective behaviors, showing how local clustering can maintain cooperation islands even when global conditions favor defection [58, 59, 60]. However, these advances remain limited by their focus on single behavioral interventions and their reliance on mean-field or simplified network approaches that miss the rich spatial dynamics arising from local clustering and cross-behavioral interactions. Furthermore, existing frameworks inadequately capture the coupling between multiple health behaviors simultaneously and fail to incorporate the critical cross-infection mechanisms that determine boundary permeability between behavioral domains [61]. This gap calls for integrated spatial models. They should track multiple health states and behavioral strategies and represent the evolutionary dynamics that drive strategy adoption, pattern formation, and stable behavioral clusters in heterogeneous populations.

We propose a lattice SIS model that fuses transmission dynamics with evolutionary strategy updating. Building on work showing that evolutionary game theory captures voluntary isolation decisions and the resulting infection waves [62], we extend the framework. Our model adds explicit spatial structure and cross-behavioral interactions that earlier models did not fully address. While earlier approaches successfully identified the feedback mechanisms between risk perception and strategy adoption in well-mixed populations, they could not capture the spatial clustering phenomena and boundary effects that prove crucial for understanding cooperation emergence in structured environments. Each agent belongs to one of four coupled states—susceptible-isolated (*S*_*Q*_), susceptible-non-isolated (*S*_*N*_), infected-isolated (*I*_*Q*_), or infected-non-isolated (*I*_*N*_)—and interacts only with its nearest neighbors. The key innovation is a cross-infection probability *p*_*a*_ that interpolates continuously between the low risk of strict isolation (*p*_*Q*_) and the high risk of unrestricted contact (*p*_*N*_). This cross-infection coupling mechanism addresses a critical limitation identified in previous evolutionary epidemiological models, where the assumption of uniform mixing obscured how behavioral boundaries can become permeable or impermeable depending on local conditions [62]. This single coupling term controls how effectively infection crosses from non-isolating to isolating contacts and, therefore, how strongly the fates of the two behavior groups are linked.

Our central objective is to chart the conditions under which cooperation can (i) shift a defecting population toward cooperation, (ii) yield persistent spatial patterning in infection and compartments, and (iii) withstand shocks to epidemiological or behavioral parameters. While previous work revealed that recurrent infection waves emerge from awareness cycles in evolutionary epidemic models, our spatial framework investigates whether these dynamics persist when agents interact locally rather than globally, while strategy payoffs respond to a shared global prevalence signal, and how spatial pattern formation can either amplify or dampen the oscillatory behaviors observed in mean-field treatments [62]. The fundamental question driving this research is: Under what conditions can cooperative isolation behaviors emerge and persist in spatially structured populations facing infectious disease threats? This encompasses identifying parameter regimes where cooperation can invade defection-dominated populations, understanding spatial patterns that facilitate cooperative spread, and determining the robustness of cooperative equilibria to perturbations. By systematically exploring the four-dimensional parameter space (*p*_*N*_, *γ, δ*, Ω)—governing transmission, recovery, perceived infection cost, and isolation cost, respectively—we uncover a rich tapestry of phases separated by sharp, often discontinuous transitions. Small rises in *p*_*a*_ switch the lattice from endemic defection to near-universal cooperation, whereas modest isolation costs can still trigger cooperative collapse. Through comprehensive parameter sweeps and spatial autocorrelation analysis using the Moran’s I index [63], we identify critical thresholds and map phase boundaries separating distinct dynamical regimes.

Beyond its epidemiological relevance, our model illuminates broader questions in nonlinear science and evolutionary game theory. We show how contact structure shapes spatial organization in infection and compartments even when behavioral switching is driven by a common global signal, clarifying when protective behavior can be sustained. Cooperative isolation is neither a fragile anomaly nor a guaranteed outcome; it is an emergent phase whose existence hinges on the delicate balance between individual incentives and collective benefits in coupled social-biological systems. From a theoretical perspective, our findings provide mechanistic insights into the nonlinear dynamics governing voluntary protective behaviors during epidemics. By identifying theoretical parameter combinations that promote cooperative isolation and critical thresholds where such behaviors collapse, we reveal fundamental principles governing behavioral-epidemic coupling that could inform future empirical studies and model development. The spatial nature of our results reveals the complex trade-offs inherent in epidemic-behavioral systems. Cooperation emergence and epidemic control represent distinct and sometimes conflicting objectives. This points toward the need for more nuanced understanding of behavioral intervention effects. This work establishes a new framework for understanding the co-evolution of disease and behavior in structured populations. It reveals fundamental principles governing collective responses to shared threats. The results demonstrate complex, sometimes paradoxical relationships between behavioral coordination and epidemiological outcomes in an interconnected world.

## 2. Methods

### 2.1. Spatially coupled epidemic-behavioral framework

We construct a spatially explicit agent-based model that integrates SIS epidemic dynamics with evolutionary game-theoretic behavioral adaptation on a structured population. Players are arranged on a *L* × *L* square lattice with periodic boundary conditions. We employ system sizes ranging from *L* = 100 to *L* = 400 to ensure robust statistics while avoiding finite-size effects. Each lattice site *i* is occupied by a single agent characterized by two coupled attributes: a health state *h*_*i*_(*t*) ∈ {*S, I* } (susceptible or infected) and a behavioral strategy *s*_*i*_(*t*) ∈ {*Q, N* } (isolation or non-isolation). This dual classification generates four distinct compartments: susceptible-isolated (*S*_*Q*_), susceptible-non-isolated (*S*_*N*_), infected-isolated (*I*_*Q*_), and infected-non-isolated (*I*_*N*_). Important terminological clarification: Throughout this manuscript, we use the subscript *Q* to denote we use “isolation” to refer exclusively to voluntary separation, in contrast to quarantine, which denotes mandatory separation of exposed individuals. Following standard public health definitions, isolation refers to voluntary separation to reduce infection transmission risk. Quarantine refers to mandatory separation of potentially exposed individuals. Our *Q* notation represents agents choosing voluntary isolation regardless of their health status. Time advances in synchronous steps so the entire lattice moves from **X**(*t*) to **X**(*t* + 1) in a single sweep. Agents interact only with their four von Neumann neighbors—for an agent at lattice position (*i, j*), these are the agents at positions (*i* + 1, *j*), (*i* − 1, *j*), (*i, j* + 1), and (*i, j* − 1), representing the four nearest neighbors in cardinal directions. This choice captures local disease spread while maintaining computational tractability.

### 2.2. Cross-behavioral epidemic transmission

Disease propagation occurs through direct contact between infected and susceptible agents on adjacent lattice sites. The transmission probability per time step depends critically on the behavioral composition of interacting pairs, reflecting the empirical observation that isolation effectiveness is determined not only by individual compliance but also by the local behavioral environment [20, 64]. For an edge connecting an infected agent of type *Y* ∈ {*I*_*Q*_, *I*_*N*_ } with a susceptible neighbor of type *X* ∈ {*S*_*Q*_, *S*_*N*_ }, the per-contact transmission probability follows the matrix:

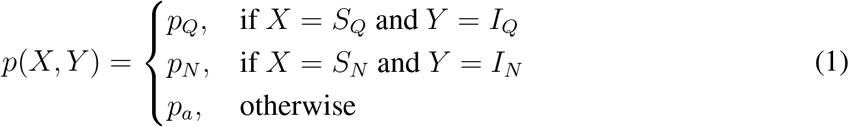

The parameter hierarchy *p*_*Q*_ *< p*_*a*_ *< p*_*N*_ reflects increasing transmission risk across behavioral boundaries. The cross–infection probability *p*_*a*_ controls how strongly defectors endanger isolated neighbors. When *p*_*a*_ ≈ *p*_*Q*_, isolation strategies maintain robust protection even against non-compliant neighbors, whereas *p*_*a*_ ≈ *p*_*N*_ renders isolation vulnerable to exploitation by defecting strategies.

All transmission events on distinct edges occur independently with probabilities determined by Eq. 1. Infected agents recover to their corresponding susceptible state with probability *γ* per time step, preserving the SIS structure that permits reinfection and sustains endemic equilibria. This recovery process maintains behavioral memory, ensuring that recovered agents retain their strategic choices.

### 2.3. Evolutionary game dynamics

We model isolation adoption as a costly cooperative strategy within a spatial public goods framework. Individual payoffs capture the fundamental trade-off between infection risk reduction and economic costs of behavioral compliance. An agent *i* employing strategy *σ*_*i*_ ∈ {*Q, N* } receives per-time-step payoff:

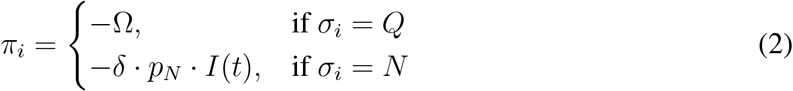

The isolation payoff − Ω represents fixed isolation costs encompassing lost income, social deprivation, and psychological stress. The non-isolation payoff − *δ*· *p*_*N*_· *I*(*t*) scales linearly with global infection prevalence *I*(*t*), where parameter *δ* converts infection probability into perceived utility loss. At the simulation initialization (*t* = 0), each agent is randomly assigned either isolation (*Q*) or non-isolation (*N*) strategy through independent Bernoulli sampling with probability *ρ* = 0.5, ensuring that exactly half the population begins with isolation behavior and half with non-isolation behavior. This random initial assignment provides a spatially mixed baseline for subsequent dynamics through the evolutionary dynamics described below, independent of any initial spatial bias toward particular strategies. While our payoff structure may appear to lack explicit returns for isolation adoption, the benefits are realized implicitly through reduced infection risk. Agents adopting isolation behavior receive protection through lower transmission probabilities and contribute to population-level infection reduction that benefits all agents. This represents a form of public goods game where the “return” manifests as risk reduction rather than direct utility gains. These benefits arise from contact-mediated transmission on the grid, which lowers exposure for agents using *Q* and, in aggregate, reduces prevalence.

This payoff formulation represents a simplification where agents of the same behavioral strategy receive identical utilities regardless of their current health state. In reality, infected individuals likely experience additional costs beyond the infection risk captured in our model, such as direct illness burden, treatment expenses, and reduced productivity. Similarly, infected agents may have different incentives for isolation adoption compared to susceptible individuals making preemptive decisions. Our simplified structure focuses on the strategic decision-making process based on population-level infection risk and isolation costs, abstracting away from health-state-dependent utility differences. This assumption enables clear analysis of epidemic-behavioral coupling mechanisms while representing a limitation for capturing the full complexity of individual decision-making during infectious disease outbreaks.

At each time step, agents revise their own strategy using a Fermi rule based on the payoff difference between the two strategies computed from *I*(*t*) (Eq. 2). Let *π*_*Q*_ = −Ω and *π*_*N*_ = −*δ p*_*N*_ *I*(*t*). Then the switching probabilities are

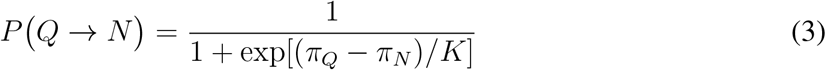

Parameter *K* controls decision-making noise, where *K* → 0 corresponds to deterministic adoption of superior strategies while *K* → ∞ represents random strategy switching [65]. This choice rule preserves computational tractability while avoiding explicit neighbor imitation on strategies; alternative social-learning mechanisms can be explored in future work.

### 2.4. Simulation protocol and parameter space exploration

Within each discrete time step, we execute updates in a fixed sequence to ensure causal consistency. First, we calculate payoffs for all agents using the current global infection prevalence *I*(*t*) measured before any state changes. Second, we perform strategy revision in parallel across all agents based on these calculated payoffs. Third, we apply disease transmission on all susceptible-infected edges using the updated behavioral strategies from the revision phase. Finally, we implement recovery events for all infected agents independently with probability *γ*.

Our parameter ranges are designed to capture some common epidemiological scenarios while ensuring comprehensive coverage of qualitatively different dynamical regimes. The transmission probability range *p*_*N*_ ∈ [0.01, 0.41] corresponds to basic reproduction numbers *R*_0_ spanning from well below to well above the epidemic threshold, encompassing scenarios from highly controlled outbreaks to rapidly spreading infections like early COVID-19 variants. The recovery probability range *γ* ∈ [0, 0.20] represents recovery timescales from 5 time steps (*γ* = 0.20) to indefinite persistence (*γ* = 0), covering acute infections like influenza to chronic conditions with slow clearance. The perceived infection cost range *δ* ∈ [0, 50] captures the spectrum from negligible risk perception (asymptomatic infections) to high-consequence scenarios (severe disease with substantial mortality or morbidity costs). The critical threshold *δ*^∗^ ≈ 5 that emerges from our analysis suggests that meaningful behavioral responses require infection costs roughly 5-10 times the baseline transmission probability, consistent with risk perception studies showing that individuals require substantial perceived threat levels to adopt costly protective behaviors. The isolation cost range Ω ∈ [0, 25] reflects economic and social isolation burdens spanning from minimal inconvenience to significant financial hardship, with the observed critical threshold Ω^∗^ ≈ 10 aligning with behavioral economics findings that cooperation collapses when individual costs exceed moderate thresholds relative to perceived benefits. While these ranges are chosen to ensure computational tractability and systematic exploration of the four-dimensional parameter space, they encompass epidemiologically relevant scenarios and reveal critical thresholds that provide theoretical insights into behavioral intervention design. Our approach prioritizes mechanistic understanding over precise quantitative calibration, establishing a framework that can be adapted and refined for specific epidemiological contexts through empirical parameter estimation. We emphasize that our parameter ranges and theoretical thresholds (such as *δ*^∗^ ≈ 5) represent dimensionless quantities within our simplified modeling framework and are not directly translatable to real-world epidemiological or behavioral measurements. Our approach prioritizes mechanistic understanding of epidemic-behavioral coupling over empirical calibration, establishing a theoretical foundation that requires substantial empirical validation and refinement before informing practical interventions.

Each simulation initializes from a random configuration characterized by initial infection density *ε* = 0.01 and cooperation fraction *ρ* = 0.5. Health states are assigned according to *h*_*i*_(0) ∼ Bernoulli(*ε*) for initial infection while strategies follow *s*_*i*_(0) ∼ Bernoulli(*ρ*) for initial cooperation, ensuring statistical independence across agents. We conduct a comprehensive exploration of the four-dimensional parameter manifold ℳ = [*p*_*N*,min_, *p*_*N*,max_] × [0, *γ*_max_] × [0, *δ*_max_] × [0, Ω_max_] through an 11^4^ = 14,641 grid design with uniform discretization intervals. The parameter ranges span transmission probability *p*_*N*_ ∈ [0.01, 0.41], recovery probability *γ* ∈ [0, 0.20], perceived infection cost *δ* ∈ [0, 50], and isolation cost Ω ∈ [0, 25]. We maintain fixed auxiliary parameters *p*_*Q*_ = 0.01 and *K* = 0.1 throughout all simulations. The cross-infection coupling strength follows 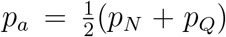, preserving the intermediate risk assumption while enabling systematic investigation of coupling effects. For each point (*p*_*N*_, *γ, δ*, Ω) ∈ ℳ, we perform 30 independent realizations to ensure statistical robustness, with temporal evolution continuing until equilibrium detection.

### 2.5. Equilibrium detection and spatial pattern analysis

The system reaches equilibrium at the first time when 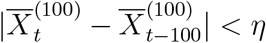 for all compartments *X* ∈ {*S*_*Q*_, *S*_*N*_, *I*_*Q*_, *I*_*N*_}, where 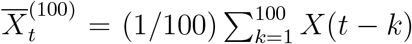 represents the 100-step moving average and *η* = 5 × 10^−4^ is the convergence tolerance. To quantify emergent spatial organization, we compute Moran’s *I* spatial autocorrelation index for each compartment:

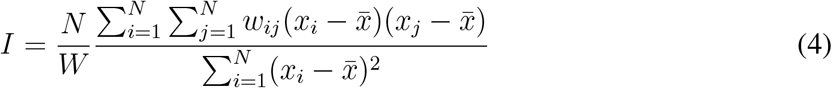

where *N* is the total number of agents, *x*_*i*_ indicates agent type (unity for target compartment, zero otherwise), 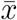 denotes the spatial mean, *w*_*ij*_ represents spatial weights (unity for adjacent pairs under von Neumann neighborhood, zero otherwise with *w*_*ii*_ = 0), and 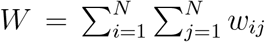 is the sum of all spatial weights. Values near zero indicate random spatial distribution while positive values reveal clustering of like strategies.

For each simulation, we record equilibrium fractions of all four compartments, complete temporal trajectories of infection prevalence *I*(*t*) and cooperation level *Q*(*t*) = *S*_*Q*_(*t*) + *I*_*Q*_(*t*), and spatial autocorrelation indices capturing pattern formation dynamics. We aggregate results across realizations and visualize them with phase diagrams, temporal trajectories, and spatial autocorrelation to identify phase boundaries, critical transitions, and emergent spatial structures.

## 3. Results

We systematically explore the four-dimensional parameter space (*p*_*N*_, *γ, δ*, Ω) using Monte Carlo simulations on 200 × 200 lattices with periodic boundaries. Unless otherwise specified, every run starts from 1% infection, 50% cooperators and selection noise *K* = 0.1. These parameter choices ensure statistical robustness while capturing the essential nonlinear dynamics of epidemic-behavioral coupling across the complete parameter space.

### 3.1. Complete phase diagram reveals rich equilibrium landscape

Figure 1 presents the complete equilibrium phase diagram across our four-dimensional parameter space, where each cell encodes the asymptotic dominant configuration among the four coupled compartments *S*_*Q*_, *S*_*N*_, *I*_*Q*_, and *I*_*N*_. Despite the theoretical possibility of 2^4^ = 16 distinct equilibrium combinations, only eight robust stationary solutions emerge consistently across independent realizations. In our parameter regime, spatial structure and cross-behavioral interactions constrain the system, eliminating configurations that might persist under mean-field assumptions. While this pattern suggests fundamental constraints imposed by spatial dynamics, our findings are limited to the specific parameter ranges investigated and cannot be generalized without broader exploration. The parameter space exhibits clear organizational principles: regions of low recovery probability (*γ* ≲ 0.04) combined with high infection cost (*δ* ≳ 15) systematically drive populations toward cooperation-dominated regimes (turquoise regions), while reverse parameter combinations favor defection-dominated attractors (orange regions).

**Figure 1.**
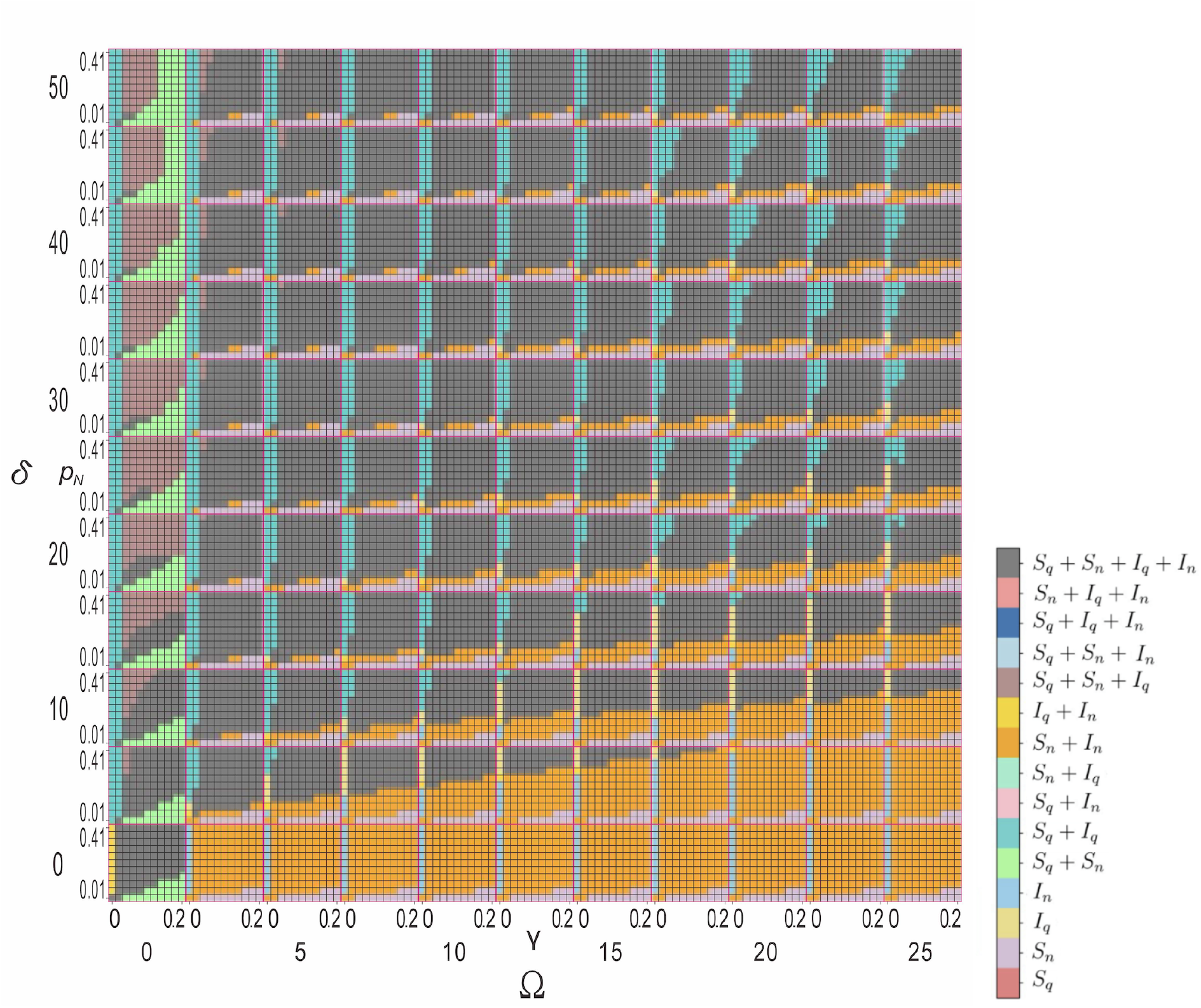
Complete phase diagram reveals eight distinct equilibrium regimes across fourdimensional parameter space. Each mini-panel displays stationary solutions for transmission probability *p*_*N*_ ∈ [0.01, 0.41], and recovery probability *γ* ∈ [0, 0.20] intervals. Columns (rows) show different values of isolation cost Ω = 0, 2.5, …, 25 (infection cost *δ* = 0, 5, …, 50), as indicated. The color code of different solutions is shown on the right. Accordingly, turquoise indicates cooperation-dominated regimes, orange marks regions where defection prevails, grey denotes fourcompartment coexistence, and blue represents susceptible-dominated states. Those regions where mixed strategies coexist are marked by intermediate colors. The complete 11^4^ parameter sweep maps stationary states across epidemiological and behavioral parameter space through systematic grid exploration.

The eight eliminated equilibrium combinations fail to persist due to fundamental incompatibilities between spatial clustering, epidemic dynamics, and behavioral incentives. Configurations dominated by infected compartments (*I*_*Q*_ or *I*_*N*_) cannot maintain stability because recovery processes systematically deplete infected populations faster than transmission can replenish them, driving the system toward susceptible-dominated states. Mixed equilibria requiring simultaneous high fractions of both isolated and non-isolated infected agents (*I*_*Q*_ + *I*_*N*_ ≫ *S*_*Q*_ + *S*_*N*_) prove unstable because differential transmission probability create evolutionary pressure favoring the more successful behavioral strategy, leading to behavioral homogenization.

Sharp phase lines cleave the diagram into qualitatively different steady states with pixel-level precision. When *δ* exceeds the critical threshold *δ*^∗^ ≈ 5 with sufficiently high *γ* and *p*_*N*_, a prominent mixed-state region emerges (grey), indicating stable four-compartment coexistence. This coexistence phase reflects a delicate balance: competing behavioral strategies persist alongside endemic infection, showing that intermediate parameters can sustain diversity via spatial pattern formation. The existence of this four-compartment equilibrium requires a precise coordination between epidemiological forcing (sufficient transmission and recovery to maintain infection circulation) and behavioral incentives (balanced costs that prevent either strategy from achieving complete dominance), revealing the intricate parameter fine-tuning necessary for maintaining stable behavioral heterogeneity in spatially structured populations. In summary, within our parameter exploration, spatial structure dramatically constrains the equilibrium landscape, reducing 16 theoretical possibilities to 8 robust configurations observed across our simulation regime. Cooperation-dominated regimes require low recovery probability (*γ* ≲ 0.04) and high infection costs (*δ* ≳ 15), establishing theoretical parameter relationships for cooperative isolation emergence within our modeling framework in spatially structured populations.

### 3.2. Dimensional reduction exposes fundamental mechanisms

To isolate the underlying mechanisms driving these complex transitions, we project the fourcompartment dynamics onto reduced dimensional subspaces corresponding to classical epidemiological and game-theoretic perspectives. Figure 2 aggregates compartments into total susceptible (*S* = *S*_*Q*_ +*S*_*N*_) and infected (*I* = *I*_*Q*_ +*I*_*N*_) populations, revealing pure epidemic dynamics stripped of behavioral heterogeneity. We emphasize that disease-free equilibria refer exclusively to states with zero infection, while the various colored regions in our phase diagrams represent different endemic equilibria with varying but non-zero infection levels.

**Figure 2.**
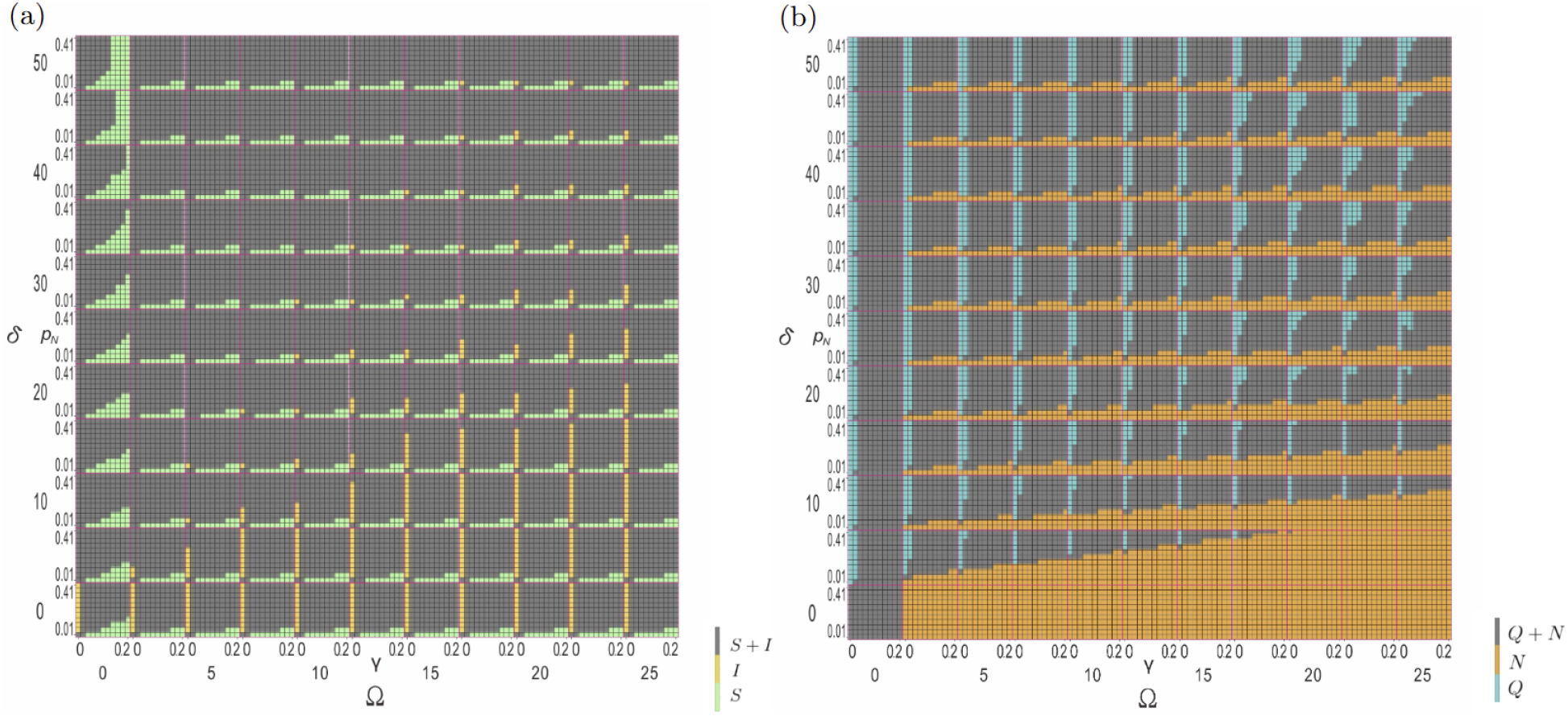
Epidemiological and behavioral projections reveal distinct organizing principles. (a) Aggregating compartments into susceptible (*S*) and infected (*I*) populations isolates disease dynamics from behavioral complexity. High prevalence (yellow) emerges exclusively when *γ* = 0; any positive recovery provides a systematic pathway toward reduced infection levels. The ultimate outcome depends on transmission intensity.. Low transmission combined with minimal isolation costs maintains susceptible-dominated states (green) even under weak recovery. (b) isolation (*Q*) versus non-isolation (*N*) populations isolate strategic dynamics from epidemiological details. Universal defection (orange) dominates when *δ* ≤ 5. Cooperation stripes (turquoise) emerge beyond this threshold, systematically narrowing with increasing isolation costs Ω until complete elimination above critical values.

The *S*–*I* projection (Figure 2 (a)) shows that sustained infection appears only when recovery is absent, that is *γ* = 0. Any positive recovery probability *γ >* 0 provides a systematic pathway toward reduced infection levels. The ultimate outcome depends on transmission intensity. Low-endemic equilibria persist under moderate transmission probability. True disease elimination occurs only when transmission probability fall below the critical threshold *p*_*N*_ ≤ 0.04. This finding establishes the primacy of biological recovery processes in determining ultimate epidemic outcomes, while behavioral factors modulate the specific dynamical trajectories toward disease elimination rather than the final equilibrium destinations. When transmission probability fall below the critical threshold *p*_*N*_ ≤ 0.04, the system converges to genuine disease-free equilibria where infection is completely eliminated and populations consist entirely of susceptible agents. This demonstrates that sufficiently weak transmission cannot sustain any level of endemic infection regardless of recovery or behavioral parameters.

Conversely, the behavioral projection (Figure 2 (b)) aggregates populations into isolation (*Q* = *S*_*Q*_ + *I*_*Q*_) and non-isolation (*N* = *S*_*N*_ + *I*_*N*_) groups, isolating the strategic decision-making dynamics from epidemiological details. The behavioral landscape reveals stark dichotomies driven by perceived infection costs. For *δ* ≤ 5, uniform orange coloration indicates near-universal defection, reflecting rational individual responses to negligible perceived risks. Beyond this critical threshold, distinct cooperation stripes emerge systematically, initially at low recovery probability and propagating upward with increasing *δ* values.

The cooperation stripes offer quantitative targets for policies that aim to trigger voluntary isolation. The critical infection cost threshold *δ*^∗^ ≈ 5 represents the minimum risk perception required to trigger the onset of cooperative behaviors, establishing a fundamental phase boundary between defection-dominated and cooperation-accessible regimes. The systematic narrowing of cooperation regions with increasing isolation costs Ω demonstrates the extreme fragility of protective behaviors under economic pressure, revealing how financial burdens can catastrophically erode voluntary compliance. Conversely, when infection costs exceed *δ* ≳ 20, cooperation regions exhibit systematic widening, indicating that sufficiently elevated risk perception can overcome economic barriers and stabilize protective behaviors even under substantial isolation costs. This asymmetric response to cost parameters—where cooperation shrinks rapidly under economic pressure but expands robustly under heightened risk perception—exposes the fundamental asymmetry in behavioral incentive structures governing epidemic response strategies. The dimensional projections reveal complementary organizing principles. Biological recovery processes determine whether systems progress toward disease elimination or maintain endemic infection. True disease-free states are achievable only under low transmission conditions regardless of behavioral composition. Meanwhile, behavioral dynamics exhibit critical threshold effects at *δ*^∗^ ≈ 5. Cooperation regions show asymmetric sensitivity to economic pressure versus risk perception, highlighting that effective interventions must address both epidemiological parameters and behavioral cost structures.

### 3.3. Critical phenomena and phase transitions

Single-parameter sensitivity analysis reveals the precise dynamical mechanisms underlying the observed phase transitions through systematic parameter variation experiments. Figure 3 (a) demonstrates transmission probability effects by varying *p*_*N*_ from 0.01 to 0.41 in increments of 0.02 while maintaining fixed control parameters *γ* = 0.02, *δ* = 25, and Ω = 20 showing stationary states of 10,000 agents. A sharp bifurcation threshold emerges at 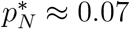, separating fundamentally distinct dynamical regimes with qualitatively different long-term behaviors. Below this critical value, the system maintains near-complete residence in the defection-dominated state (*S*_*N*_ + *I*_*N*_ *≈* 1), reflecting insufficient epidemiological selection pressure to overcome isolation adoption costs and trigger cooperative transitions.

**Figure 3.**
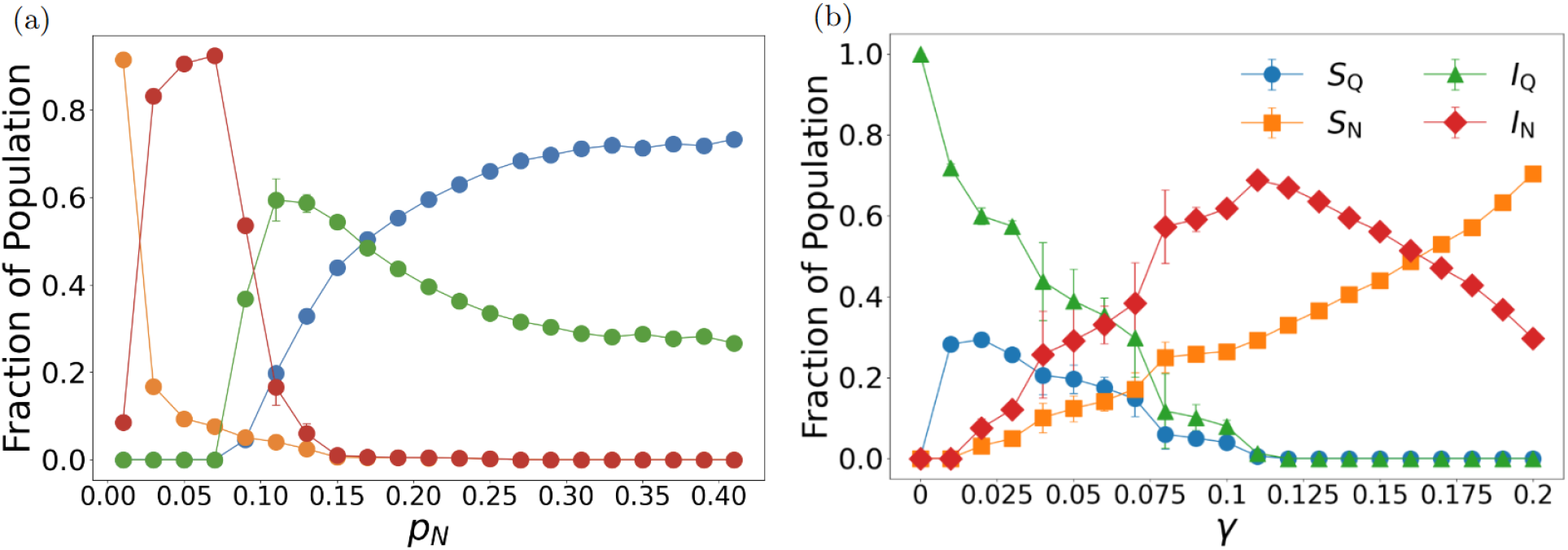
Critical thresholds govern abrupt behavioral transitions through nonlinear feedback mechanisms. Compartment fractions showing stationary states of 10,000 agents with error bars representing standard deviations from 30 independent experiments. (a) Transmission probability dependence with fixed parameters *γ* = 0.02, *δ* = 25, and Ω = 20. Critical transition at 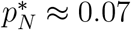 separates low-cooperation from high-cooperation regimes, with secondary transition at 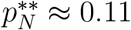 initiating protected susceptible regime. The *I*_*N*_ peak during first transition demonstrates rapid collective behavioral switching, while the inverse relationship between transmission probability and total infection beyond 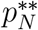 reveals nonlinear feedback mechanisms governing voluntary epidemic control. (b) Recovery probability dependence with fixed parameters *p*_*N*_ = 0.09, *δ* = 30, and Ω = 17.5. Increasing *γ* systematically reduces infection and cooperation through diminished selection pressure. Narrow *I*_*N*_ peak near *γ*^∗^ ≈ 0.11 reflects delicate balance between defection temptation and infection control, revealing nonlinear interactions between biological and behavioral timescales.

The transition at 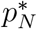 triggers a dramatic behavioral cascade characterized by rapid collective strategy switching through spatial contagion effects. Once *p*_*N*_ exceeds this threshold of approximately 0.07, the infected non-isolation fraction *I*_*N*_ rises sharply in a characteristic transient spike before being systematically displaced by a dramatic surge in infected isolation adoption *I*_*Q*_. This sharp rise and subsequent decline of *I*_*N*_ signals population-wide recognition of elevated infection risks and initiation of protective behavioral responses. The high perceived infection cost (*δ* = 25) relative to the low recovery probability (*γ* = 0.02) drives this rapid switch to cooperative behavior, demonstrating how infinitesimal increases in transmission intensity can trigger discontinuous cooperative phase transitions when individual cost-benefit calculations cross critical decision thresholds.

Beyond a secondary transition point 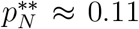, the system enters a “protected susceptible” regime where *S*_*Q*_ achieves dominance in population composition while overall infection prevalence decreases monotonically with increasing *p*_*N*_. This creates a paradoxical inverse relationship: higher transmission potential generates lower actual infection levels. This emerges from nonlinear feedback between transmission risk and behavioral adaptation. Elevated transmission threats trigger preemptive isolation adoption that more than compensates for increased infection pressure, creating a negative feedback loop that enhances population protection. This outcome represents successful epidemic control through voluntary behavioral adaptation achieved without external enforcement mechanisms, constituting a striking manifestation of self-organized collective protection through evolutionary game dynamics.

Recovery probability sensitivity analysis reveals equally dramatic threshold phenomena with distinct characteristics (Figure 3 (b)). We varied *γ* from 0 to 0.2 in increments of 0.01 while maintaining fixed parameters *p*_*N*_ = 0.09, *δ* = 30, and Ω = 17.5 over 10^4^ time steps, analyzing stationary states of 10,000 agents. At low recovery probability (*γ* ≲ 0.025), persistent infection sustains elevated cooperation levels with high *I*_*Q*_ and *S*_*Q*_ fractions through continuous threat perception and maintained selection pressure favoring protective behaviors. Increasing recovery probability systematically reduce both infection prevalence and cooperation incentives, creating a monotonic decline in isolation adoption as disease threats diminish and reduced epidemiological pressure weakens the incentive for self-isolation.

A narrow *I*_*N*_ peak at *γ* ≈ 0.11 marks the point where recovery is quick enough to tempt defection but still slow enough for flare-ups. This peak reveals a critical transition zone where recovery becomes fast enough to tempt defection but remains insufficient to prevent infection outbreaks, demonstrating complex nonlinear interactions between epidemiological and behavioral timescales. The emergence of this intermediate regime produces counterintuitive outcomes that deviate from monotonic expectations, highlighting the non-trivial coupling between biological recovery processes and strategic behavioral responses. The single-parameter sensitivity analysis reveals sharp critical transitions at approximately 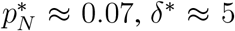, and Ω^∗^ ≈ 10. These transitions demonstrate nonlinear feedback mechanisms where infinitesimal parameter changes trigger discontinuous behavioral cascades and paradoxical outcomes such as voluntary epidemic control, revealing the theoretical complexity of epidemic-behavioral coupling.

Behavioral parameter sensitivity confirms the critical role of risk perception in driving evolutionary transitions. We varied *δ* from 0 to 25 in unit steps while maintaining fixed parameters *p*_*N*_ = 0.41, *γ* = 0.04, and Ω = 22.5, analyzing stationary states of 10,000 agents. Infection cost analysis (Figure 4 (a)) reveals a characteristic two-stage transition initiating at *δ*^∗^ ≈ 5. For *δ <* 5, the penalty of infection remains trivial relative to isolation costs, resulting in dominance by non-isolated infected agents (*I*_*N*_). Initial isolation adoption occurs preferentially among infected individuals as *δ* rises above 5 (evidenced by the *I*_*Q*_ surge peaking near *δ*^∗∗^ ≈ 7), followed by preemptive adoption among susceptible populations (demonstrated by subsequent *S*_*Q*_ growth that gradually replaces *I*_*Q*_). This indicates that higher perceived infection costs drive more agents to adopt isolation, thereby reducing overall infection levels and establishing a stable, low-infection, high-isolation equilibrium.

**Figure 4.**
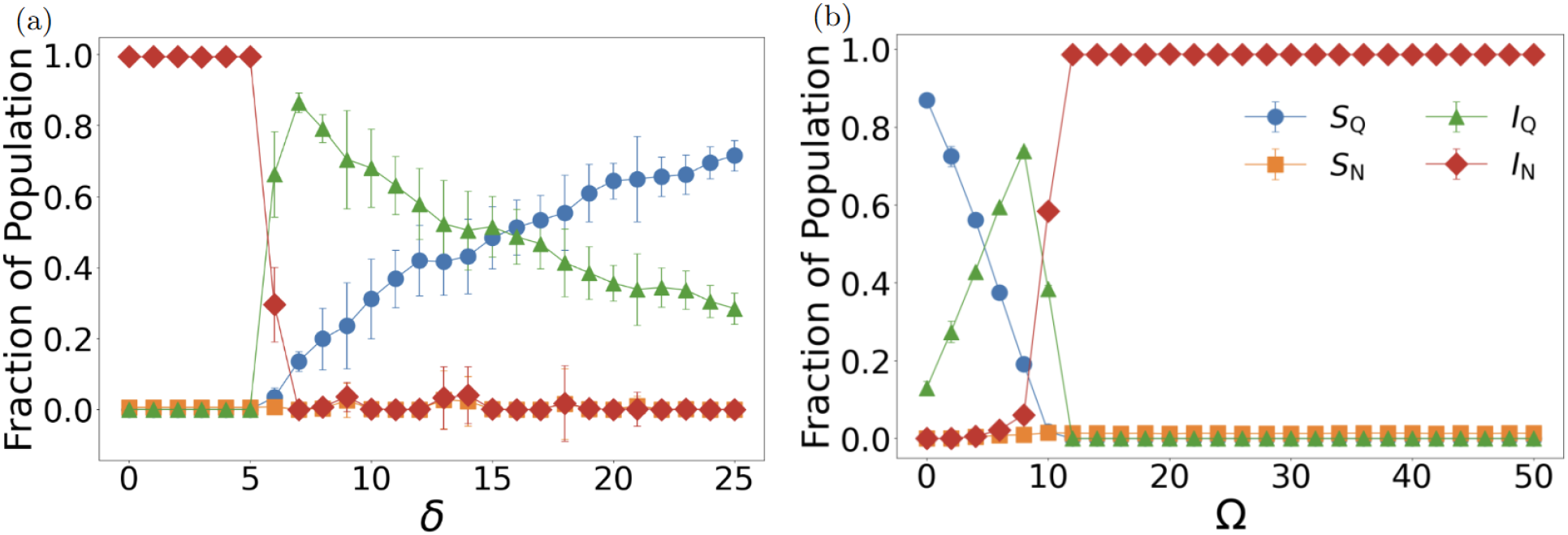
Behavioral parameter sensitivity reveals critical thresholds for cooperation emergence and collapse. Compartment fractions showing stationary states of 10,000 agents with error bars representing standard deviations from 30 independent experiments. (a) Risk perception dependence with fixed parameters *p*_*N*_ = 0.41, *γ* = 0.04, and Ω = 22.5. Transition at *δ*^∗^ ≈ 5 initiates two-stage isolation adoption: initial adoption among infected populations (*I*_*Q*_ peak around *δ* ≈ 7) followed by preemptive adoption among susceptibles (*S*_*Q*_ growth). Below threshold, infection penalties remain trivial relative to isolation costs, resulting in *I*_*N*_ dominance. Beyond the peak, cooperative susceptibles accumulate as prevalence declines, establishing stable low-infection, highisolation equilibrium. (b) Economic pressure dependence with fixed parameters *p*_*N*_ = 0.21, *γ* = 0.02, and *δ* = 5. Low costs (Ω *<* 4) sustain mixed states with coexisting isolated and non-isolated populations. Sharp transition at Ω^∗^ ≈ 10 eliminates both isolated fractions simultaneously while *I*_*N*_ climbs, demonstrating brittleness of cooperative equilibria under economic pressure. This critical threshold demonstrates theoretical relationships between cost parameters and behavioral transitions.

Lastly, we also varied isolation costs Ω from 0 to 50 in increments of 2 while maintaining fixed parameters *p*_*N*_ = 0.21, *γ* = 0.02, and *δ* = 5. The isolation cost analysis (Figure 4 (b)) exposes extreme sensitivity to economic intervention burdens. At low isolation costs (Ω *<* 4), moderate levels of isolated fractions are maintained in a mixed state. A sharp collapse transition at Ω^∗^ ≈ 10 triggers simultaneous elimination of both isolated fractions while *I*_*N*_ rises sharply, revealing the inherent brittleness of cooperative equilibria under economic pressure. This trend demonstrates that higher perceived isolation costs discourage cooperative behavior, leading the system toward a state dominated by non-isolated agents. This critical threshold provides theoretical relationships relevant to behavioral dynamics: maintaining intervention costs below Ω^∗^ emerges as essential for preserving voluntary compliance and avoiding catastrophic cooperation collapse.

### 3.4. Cross-infection coupling as master control parameter

The cross-infection probability *p*_*a*_ emerges as the fundamental control parameter determining system-wide evolutionary outcomes. Figure 5 presents three representative scenarios spanning *p*_*a*_ ∈ {0.01, 0.05, 0.10 }, obtained from single *N* = 10,000 agent runs lasting 10,000 time steps with fixed parameters Ω = 6, initial cooperation fraction 50%, initial infection fraction *ε* = 0.01, recovery probability *γ* = 0.05, and *δ* = 10, with associated transmission probabilities *p*_*N*_ = 1.5 *p*_*a*_ and *p*_*Q*_ = 0.5 *p*_*a*_, maintaining the desired ratio *p*_*N*_ : *p*_*a*_ : *p*_*Q*_ = 3 : 2 : 1.

**Figure 5.**
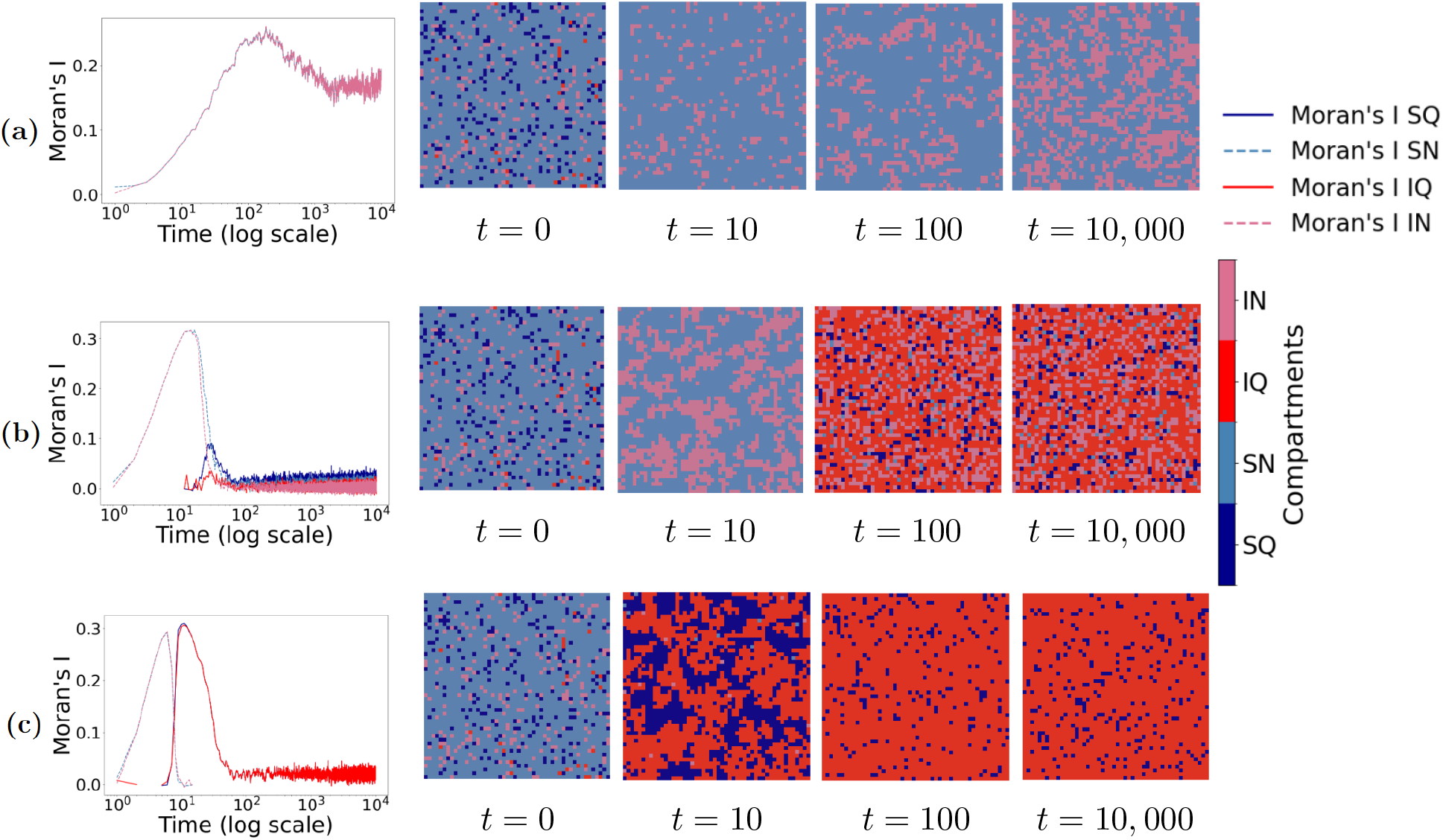
Cross-infection coupling controls spatiotemporal pattern formation and determines epidemic-behavioral outcomes. Spatio-temporal patterns for three cross-infection probability *p*_*a*_ = 0.01, 0.05, 0.10 obtained from single *N* = 10,000 agent runs lasting 10,000 time steps with fixed parameters Ω = 6, *γ* = 0.05, *δ* = 10, initial cooperation fraction 50%, and initial infection fraction *ε* = 0.01, with associated probability *p*_*N*_ = round(1.5 *p*_*a*_) and *p*_*Q*_ = round(0.5 *p*_*a*_). In each row, the left panel shows Moran’s *I* indices for *S*_*Q*_ (dark-blue solid), *S*_*N*_ (light-blue dashed), *I*_*Q*_ (red solid), and *I*_*N*_ (pink dashed), while the four right panels display lattice snapshots at *t* = 0, 10, 100, 10,000. (a) Low coupling (*p*_*a*_ = 0.01) eliminates isolation behaviors immediately, creating two-compartment dynamics with progressive boundary smoothing and domain coarsening. (b) Intermediate coupling (*p*_*a*_ = 0.05) sustains four-compartment coexistence through sporadic emergence of protective clusters soon after *t* ≈ 10, followed by stable spatial pattern formation. (c) High coupling (*p*_*a*_ = 0.1) drives explosive simultaneous outbreaks of isolation behaviors, leading to rapid coordination toward uniform cooperation, yet paradoxically results in persistent high infection levels as the system becomes dominated by infected-isolating agents (*I*_*Q*_).

At low coupling strength (*p*_*a*_ = 0.01), isolation populations disappear immediately, leaving only *S*_*N*_ and *I*_*N*_ compartments on the entire lattice. The low infection prevalence that results from minimal cross-infection (typically below 5% of the population) fails to create sufficient selective pressure for isolation adoption, demonstrating how weak epidemic threats paradoxically undermine the very behaviors needed for epidemic control. The system reduces to a classical two-compartment epidemic with standard spatial segregation patterns where boundaries between light red (*I*_*N*_) and light blue (*S*_*N*_) regions gradually become smoother as jagged edges are absorbed and straightened, resulting in larger coherent domains with reduced boundary lengths. Intermediate coupling (*p*_*a*_ = 0.05) reveals a fundamentally different pattern: soon after *t* ≈ 10, sporadic isolation infection clusters (*I*_*Q*_) emerge, surrounded by thin layers of isolation susceptible populations (*S*_*Q*_). This sustains all four compartments through spatial domain formation, creating complex behavioral mosaics with well-defined protective boundaries. High coupling (*p*_*a*_ = 0.10) triggers simultaneous explosive outbreaks of both infection and isolation behaviors: *S*_*Q*_ and *I*_*Q*_ fractions first drop to zero briefly, then rapidly expand into large domains, while non-isolation populations are almost completely eliminated.

Moran’s I spatial autocorrelation analysis provides quantitative insight into the underlying pattern formation dynamics across these regimes. For low coupling (*p*_*a*_ = 0.01), both *S*_*N*_ and *I*_*N*_ Moran’s I indices climb together and stabilize at elevated levels, reflecting the progressive spatial sorting and boundary smoothing that creates large, coherent domains with perfect anti-correlation. Under intermediate coupling (*p*_*a*_ = 0.05), the emergence of isolation clusters soon after *t* ≈ 10 drives sharp peaks in *I*_*Q*_ and *S*_*Q*_ autocorrelation indices while *S*_*N*_ values decline, followed by compression of all curves to low, flat levels as the four-compartment coexistence stabilizes. High coupling (*p*_*a*_ = 0.10) produces dramatic Moran’s I spikes for isolation compartments as large domains rapidly form, while non-isolation indices permanently collapse to zero. As *I*_*Q*_ clusters merge, the red curve gradually declines and stabilizes at moderate levels, corresponding to a final lattice configuration dominated almost entirely by isolation infection (*I*_*Q*_) with only scattered healthy points remaining.

These findings establish *p*_*a*_ as a coupling strength parameter governing inter-group interactions and pattern formation. The three coupling regimes correspond to fundamentally different epidemic outcomes: complete isolation failure with classical epidemic spread (*p*_*a*_ = 0.01), balanced four-compartment coexistence with localized protective clustering (*p*_*a*_ = 0.05), and near-universal isolation adoption with failed epidemic suppression (*p*_*a*_ = 0.10). Below critical coupling values, behavioral groups remain effectively isolated, preventing cooperation spread through population barriers. Above critical values, strong coupling drives universal coordination through spatial contagion of protective behaviors, demonstrating how local interactions can generate global synchronization phenomena and transform epidemic trajectories from uncontrolled spread to collective protection.

This coupling-dependent behavior reveals a fundamental paradox in epidemic-behavioral systems: the cross-contact that public health interventions typically aim to minimize is actually essential for cooperative isolation behaviors to emerge and spread. At low coupling, insufficient cross-infection creates weak selective pressure for isolation adoption, while the slow spread of infection provides inadequate opportunity for cooperative behaviors to be observed and reflected in the global prevalence signal that shifts incentives uniformly. This creates a “cooperation extinction” scenario where protective behaviors cannot gain sufficient traction to persist. At intermediate coupling, cross-infection provides sufficient threat to motivate isolation behaviors while producing contact-driven spatial patterning under a shared, prevalence-based decision signal, yet this still results in mixed behavioral states with persistent infection across all compartments. Most remarkably, the high coupling regime reveals a striking paradox. While it successfully drives near-universal isolation adoption, it paradoxically sustains the highest infection levels of all three scenarios. This demonstrates that excessive cross-contact can overwhelm the protective benefits of widespread cooperation. The system converges to a state where nearly everyone isolates yet nearly everyone remains infected, creating an IQ-dominated equilibrium. This finding challenges conventional intuitions about the relationship between cooperation and epidemic control, revealing that behavioral coordination and epidemiological success are not synonymous in strongly coupled systems.

## 4. Discussion

The coupled epidemic–behavior lattice uncovers nonlinear behavior that rises directly from the feedback between infection and strategy choice. Our exploration of the four-dimensional parameter space reveals sharp phase boundaries that divide the system into three attractors: endemic defection basins, fragile coexistence, and robust cooperation. Tiny shifts in cross infection or small cuts in isolation cost flip the lattice from widespread defection to almost full cooperation. High coupling forces nearly everyone to isolation yet still leaves a stubborn endemic core, showing that cooperation alone cannot assure disease control.

Cross-infection coupling acts as the master control: low coupling yields isolation failure and classical spread; intermediate coupling supports four-compartment coexistence with protective clustering; high coupling drives near-universal isolation with persistent endemic infection. The structures exhibit remarkable robustness against perturbations, maintaining their organizational integrity under stochastic fluctuations and parameter drift across extended time simulations. In well-mixed models, higher risk perception lowers peaks via extended isolation; in contrast, our spatial framework shows that the same coordination can sustain endemic infection by creating persistent isolation-infected clusters.

From a theoretical perspective, our results illuminate fundamental mechanisms underlying epidemic-behavioral coupling and reveal why simple intervention strategies may exhibit limited efficacy in complex adaptive systems. Our framework demonstrates that epidemic-behavioral systems exhibit counter-intuitive dynamics, such as the paradoxical requirement for intermediate cross-contact levels to sustain cooperative isolation behaviors. While these insights suggest important principles for understanding voluntary protective behaviors, translating our theoretical findings into practical interventions would require substantial model extensions, empirical calibration with real behavioral and epidemiological data, and validation across diverse population contexts. The critical thresholds we identify represent dimensionless theoretical quantities within our simplified framework rather than directly actionable policy parameters.

Our findings fundamentally challenge classical evolutionary intuitions regarding cooperation sustainability. We reveal how epidemic-behavioral coupling creates reactive fitness landscapes. In these landscapes, when more agents choose non-isolation, infection prevalence rises, which elevates the payoff for isolation and incentivizes cooperation. This mechanism extends beyond isolation behaviors to other health interventions: evolutionary models of vaccination decisions reveal similar paradoxes where individual strategy choices based on infection prevalence and intervention costs can lead to Nash equilibria that differ substantially from social optima, with waning immunity and multi-strain dynamics further complicating the strategic landscape [66, 67]. Classic theory says free riders win, but here each defector sparks local outbreaks that raise the appeal of isolation and launch waves of conversion. This mechanism provides a spatial resolution to the temporal instability problem observed in mean-field evolutionary epidemic models, where voluntary isolation strategies simultaneously reduce individual infection risk while creating conditions for their own abandonment through reduced disease awareness [17]. Spatial structure enables the formation of self-sustaining cooperative enclaves that persist independently of global epidemic conditions, explaining the striking epidemiological heterogeneity observed across demographically similar populations during real epidemic events.

The critical phenomena we observe—characterized by sharp transitions at precisely defined thresholds—suggest that epidemic-behavioral systems operate near critical points where small parameter changes drive large-scale reorganization [68, 69]. Our Moran’s I autocorrelation analysis quantifies the underlying pattern formation dynamics, revealing three characteristic phases: rapid nucleation with sharp autocorrelation peaks, competitive growth through domain boundary formation, and dynamic stabilization via plateau convergence. Such spatial signals provide theoretical insights into pattern formation dynamics. Future empirical work could investigate whether these patterns might serve as early warning indicators in real-world settings. The framework’s predictive power extends beyond individual epidemic events to illuminate fundamental principles governing voluntary collective action under shared threats across diverse domains.

The emergence of transmission-driven spatial clustering establishes epidemic-behavioral coupling as a paradigmatic example of complex adaptive systems exhibiting critical phenomena and emergent spatial organization. Through systematic exploration of our four-dimensional parameter manifold via comprehensive parameter combinations, we reveal a rich landscape of equilibrium phases separated by knife-edge boundaries that shift dramatically with single parameter increments. Critical thresholds emerge with precision—transmission thresholds, risk perception thresholds, and economic pressure thresholds—separating fundamentally different equilibrium phases. The abrupt nature of these transitions demonstrates that the system operates near critical points where small perturbations drive large-scale reorganization. Spatial autocorrelation analysis quantifies the underlying pattern formation dynamics, revealing three characteristic phases: rapid nucleation with sharp autocorrelation peaks, competitive growth through domain boundary formation, and dynamic stabilization via plateau convergence. This framework demonstrates emergent spatial organization from local interactions. Critically, it shows that cooperation emergence does not guarantee epidemic control.

While our framework provides fundamental insights into epidemic-behavioral coupling, several limitations warrant acknowledgment and suggest avenues for future research. For tractability, utilities are strategy-specific rather than health-state dependent; allowing state-contingent costs is an important extension. Additionally, our strategy updates use a prevalence-based rule; future work could compare alternative social-learning mechanisms (e.g., majority or multi-peer heuristics). Our model assumes homogeneous agents with identical payoff structures, whereas real populations exhibit substantial heterogeneity in risk perception, economic capacity, and social connectivity [44]. The assumption of perfect information about global infection prevalence represents another simplification, as individuals rely on incomplete and potentially biased information sources [59]. Moreover, our claims about equilibrium reduction from 16 to 8 possible configurations are based solely on our specific four-dimensional parameter exploration and cannot be generalized to all possible parameter regimes without additional systematic investigation. While the observed reduction suggests fundamental constraints imposed by spatial structure and epidemic-behavioral coupling, broader parameter space exploration would be needed to establish the universality of this phenomenon. Future extensions incorporating health-state-dependent payoffs, agent heterogeneity, empirical network topologies, information propagation dynamics, continuous strategy spaces, and demographic processes would enhance realism and predictive power. Experimental validation through controlled behavioral studies and empirical calibration using real epidemic data represent crucial next steps for translating theoretical insights into practical applications, while the binary strategy framework could be extended to capture multiple behavioral interventions simultaneously.

Our analysis focuses on voluntary isolation, and this coupled epidemic-behavioral framework can be generalized to other protective behaviors involving individual cost-benefit trade-offs and social influence dynamics. Behaviors such as mask wearing, hand hygiene, and testing compliance share fundamental characteristics with isolation in that they balance personal costs against infection risk reduction, but they involve distinct epidemiological mechanisms that would require framework modifications [23, 24, 58]. Unlike quarantine, which removes individuals from transmission entirely, these behaviors reduce transmissibility or susceptibility by multiplicative factors while maintaining contact [20]. This would require modification of our transmission probability matrix to include intermediate protection levels rather than binary high-risk/low-risk states. The payoff structures would similarly need adaptation to capture behavior-specific costs: mask wearing involves procurement and comfort costs with lower economic burden than isolation, while testing involves financial costs and psychological stress from potential positive results. Critically, the transmission-clustering phenomena we observe may manifest as protective patterns with more permeable boundaries for mask wearing, or temporal patterns for periodic testing behaviors. This suggests that our critical threshold framework may reveal general principles governing voluntary compliance across diverse public health interventions.

The framework established here offers conceptual insights and computational tools for understanding the fundamental challenge of voluntary collective action during epidemics, where the co-evolution of biological and social processes creates complex adaptive dynamics. While our theoretical discoveries reveal important principles governing cooperation emergence in spatially structured populations, future work must bridge the gap between these mechanistic insights and practical application through empirical validation, model calibration, and integration of real-world complexity factors that our simplified framework cannot capture.

## Data and Code Availability

The simulation data supporting the findings of this study and the source code used to generate the results are available in the GitHub repository at: https://github.com/Research-Repo123/Quarantine-Solitons-Research/tree/main. Additional data files and extended simulation results are available upon reasonable request from the corresponding author.

